# Impact of a White Matter Reference Region on the Relationship between Florbetapir PET Measurements of Amyloid Plaque Deposition and Measurements of Cognitive Decline

**DOI:** 10.1101/2025.09.19.25336123

**Authors:** V Bhargava, M Wang, Y Chen, J Luo, M Weiner, S Landau, W Jagust, M Sabbagh, Y Su, EM Reiman, K Chen

**Affiliations:** University of Arizona College of Medicine Phoenix; Banner Alzheimer’s Institute; Arizona State University; Translational Genomics Research Institute; University of California San Francisco; University of California, Berkeley; Barrow’s Neurological Institute; Arizona Alzheimer’s Consortium

**Keywords:** “Amyloid Beta Deposition”, “Cognition”, “Alzheimer’s Disease”, “Standarized Uptake Value Ratios”, “Florbetapir”, “PET”

## Abstract

The objective of this study was to systematically investigate both cross-sectional and longitudinal associations between amyloid PET tracer, Florbetapir (FBP), and cognition when different reference regions of interest – whole cerebellum versus white matter – are used for Standardized Uptake Value Ratio (SUVR) semi-quantification of amyloid beta deposition. Baseline and 2.2±0.4 year follow-up Florbetapir PET scans from 1,238 mild AD dementia, mild cognitive impairment (MCI), and cognitively unimpaired (CU) participants from AD Neuroimaging Initiative (ADNI) were used to characterize and compare the impact of using a cerebral white matter versus whole cerebellar reference region on cross-sectional and longitudinal relationships between florbetapir SUVR indicators of amyloid plaque deposition and measurements of cognitive or clinical decline (ADAS-Cog-13, CDR-Sum Boxes, and AVLT-total) after covarying for age and education. In both cross-sectional and longitudinal comparisons, florbetapir PET measurements of amyloid plaque deposition using the cerebral white matter reference region were more closely related to each measure of cognitive or clinical decline in the aggregate mild dementia, MCI and CU group (P<1.3E-06). This study supports the potential use of a cerebral white matter reference region in the detection and tracking of amyloid plaque deposition using florbetapir PET. Additional studies are needed to clarify the generalizability of findings to other amyloid PET ligands.

## Introduction

The study of Alzheimer’s disease requires accurate and standardized global PET measurements of in-vivo amyloid plaque deposition typically expressed as Standard Uptake Value Ratios (SUVRs) (Suppiah et al., 2019). For a given radiotracer, SUVRs are calculated by computing the PET signal ratio from a target cerebral cortical region-of-interest (ROI) to reference ROI that is relatively spared from amyloid pathology (Pemberton et al., 2022). While the whole cerebellum is most commonly used, other reference regions of interest include cerebellar gray matter, pons, and cerebral white matter. A whole cerebellum reference ROI is commonly used for cross-sectional measurements as well as longitudinal studies in which SUVRs are transformed into Centiloid measurements (Iaccarino et al., 2025). A cerebellar white matter reference ROI has shown greater power to track longitudinal changes in amyloid PET measurements and stronger correlations between these changes and rates of clinical decline, at least for florbetapir (Chen et al., 2015).

While in-vivo research has focused on identifying amyloid positivity in cross-sectional assessments or tracking changes in amyloid accumulation in longitudinal studies, the relationship between amyloid deposition and cognitive decline deserves more attention. Out of the few studies reported so far, most have utilized a cerebellar reference region and demonstrated inconsistent or weak correlations between amyloid deposition and cognition (Ciarmiello et al., 2019; Hanseeuw et al., 2019; Souto et al., 2021; Stevens et al., 2022; Villemagne et al., 2013; Villemagne et al., 2011). Similarly, post-mortem studies have inconsistently shown limited or no significant relationship between amyloid plaque density and dementia severity (Aizenstein et al., 2008; Guillozet et al., 2003; Michalowska et al., 2022).

We previously demonstrated improved power in tracking two-year changes in Florbetapir SUVRs for evaluating amyloid-modifying treatment effects when a cerebral white matter reference ROI is used rather than a cerebellar reference ROI (Chen et al., 2015). In this study, we systematically investigate both cross-sectional and longitudinal associations between amyloid and cognition when different reference regions – cerebellum versus white matter – are used.

## Methods

### Study Design

Alzheimer’s Disease Neuroimaging Initiative (ADNI) is a longitudinal, multisite study spanning across 57 study sites in the United States and Canada, created to track multiple clinical, genetic, blood-based, CSF and various neuroimaging biomarkers for AD. This dataset includes elderly control patients, Mild Cognitive Impairment (MCI), and AD patients followed longitudinally.

### Participants

Our study population included data from total of 1238 participants with the following distribution: between n=378 for CU (n=86 amyloid positive individuals and n=293 amyloid negative individuals), n=592 for MCI, and n=267 for AD, all underwent both FBP and FDG PET scans concurrently. A total of 6 visits from the MCI dataset and 7 visits from the AD dataset were excluded due to missing data. Full inclusion and exclusion criteria can be found at www.adni-info.org. All subjects were between the ages of 55-90 years old, provided their informed consent, and did not have any other neurological condition. MCI patients had a Clinical Dementia Rating (CDR) of 0.5 with a memory box score of at least 0.5 while AD patients had a CDR of 0.5 or 1.The general clinical/behavioral performance of MCI patients was such that an on-site diagnosis of AD could not be made by a physician. NINCDS and ADRDA criteria were followed for AD diagnosis (McKhann et al. 1984).

### FBP PET

FBP PET data was acquired using standardized ADNI protocols on various PET scanners. Data were corrected for radiation-attenuation and scatter using transmission scans or X-ray CT, and reconstructed using reconstruction algorithms specified for each type of scanner as described at www.loni.ucla.edu/ADNI/Data/ADNI_Data.shtml. Acquired images were reviewed, pre-processed, and standardized by ADNI PET Coordinating Center investigators at University of Michigan. The images were then uploaded to the Laboratory of Neuroimaging (LONI) ADNI website previously at UCLA and currently at USC, and ultimately downloaded from the LONI website in NIFTI format by investigators at the Banner Alzheimer’s Institute for the analyses in this report. Details about FBP images can be found at http://adni-info.org. FBP-PET data was acquired in 5-min frames from 50 to 70 min post-injection. FBP PET scans were acquired and pre-processed following the ADNI pipeline (http://adni.loni.usc.edu/methods/pet-analysis-method/pet-analysis/). FBP images were spatially normalized to MNI template using Statistical Parametric Mapping 12 (SPM12).

### FBP SUVRs

SPM12 was used to extract FBP uptake from mean cortical (mcROI) relative to uptake in (1) cerebral white matter regions and (2) cerebellar regions, masks of all 3 are pre-defined in the MNI template in our previous investigation (Chen et al. 2015). Mean Cortical SUVRs (mcSUVRs) were calculated by dividing FBP uptake in mean cortical ROI by FBP uptake in cerebral WM reference region of interest (mcSUVR_wm_) or by FBP uptake in a cerebellum reference region of interest (mcSUVR_cb_). The cerebral white matter reference region included a collection of voxels in the right and left corpus callosum (using the corpus callosum mask from the WFU_PickAtlas toolbox) and right and left centrum semiovale excluding voxels closest to the grey matter or to the ventricles (for more details refer to Chen et al. 2015). For the cerebellar reference ROI, tracer uptake in the whole cerebellum was used.

### Cognitive Tests

Alzheimer’s Disease Assessment Scale-Cognitive (ADAS-Cog 13): This test evaluates memory, reasoning, language, orientation, ideational praxis, and constructional praxis. Scores range from 0 (best) to 70 (worse) with higher scores indicating poorer performances. ADAS-Cog 13 includes the addition of delayed word recall and number cancellation. More information can be found in (Rosen et al., 1984).

Clinical Dementia Rating Sum Boxes (CDR-Sum Boxes): This test examines global measure of severity of dementia by testing six categories of cognitive functioning including memory, orientation, judgement/problem solving, community affairs, home and hobbies, and personal care. The ratings are then synthesized into a global rating of dementia which ranges from 0-3 with a more refined score. More information can be found in (Berg, 1988).

Rey Auditory Verbal Learning Test (AVLT Total): This test is used to assess learning and memory by providing participants with 5 learning trials. During each trial, 15 unrelated nouns are spoken at a rate of one word per second and the patient is asked to immediately recall as many words as possible. After, a 30-minute delay with unrelated testing is administered and free recall of the original 15 words is elicited. Lastly, a yes/no recognition test with 15 of the original words and 15 unrelated distractor words is administered. A version including both short term memory and long-term memory was administered (AVLT-Total). More information can be found in (Rey 1964).

### Statistical Analyses

ANOVA was used to compare group demographic characteristics and are summarized in Table 1. We used partial correlation to assess the relationship between global Aβ deposition, using the cerebellum and white matter as reference regions (mcSUVR_cb_ and mcSUVR_wm_), and measures of cognitive/memory impairment. Partial correlations were calculated covarying out baseline age and education. A cross-sectional analysis, comparing baseline associations, and a longitudinal analysis, comparing the association between change in cognitive/memory impairment and change in mcSUVRs, were each conducted.

**Table 1:** Demographics Table.

| Characteristic | CU (n=379) | MCI (n=592) | AD (n=267) |
| --- | --- | --- | --- |
| Age (years) | 75 ± 7 | 73 ± 8 | 75 ± 8 |
| Sex (n [%] female)) | 202 (54%) | 261 (43%) | 109 (42%) |
| Education (years) | 16.5 ± 2.6 | 16.1 ± 2.7 | 15.9 ± 2.6 |
| MMSE | 29.0 ± 1.3 | 28.0 ± 1.8 | 22.5 ± 3.2 |
| ADAS-Cog13 | 9.1 ± 4.6 | 15.3 ± 7.0 | 31.5 ± 9.6 |
| APOE4 Carriers (n [%]) | 97 (27%) | 242 (43%) | 117 (57%) |
| # of participants with >2 visits | 161 | 227 | 46 |

Steiger’s Z-test was then used to determine if the difference between partial correlation coefficient of cognitive or memory impairment/mcSUVR_cb_ and cognitive or memory impairment/mcSUVR_wm_ were significant, with p=0.05. Partial correlations were examined among all subjects and within each diagnostic group: AD, mild cognitive impairment (MCI), and cognitively unimpaired (CU) patients.

## Results

Subject characteristics are described in Table 1. The dataset from a total of 1238 participants was included in the cross-sectional analysis of the study with CU (n=379), MCI (n=592), and AD (n=267). All 434 participants with two or more visits, with an average time between visits of 2.16±0.37 years, were included for the longitudinal analysis with CU (n=161), MCI (n=227), and AD (n=46). Patients’ demographic characteristics including sex, education, age, number of months since baseline or visit number, and MMSE scores are shown in Table 1. Significant differences were observed between mean age (p=5.02E-05), number of females (p=0.0018), MMSE and ADAS-Cog13 scores (p=0.013 and p=4.75E-212), and number of years of education (p=3.28E-198) between AD, MCI and CU groups, respectively. Average numbers of visits for participants with longitudinal data were 2.02 for CU participants, 2.19 for MCI participants, and 2.04 for AD participants.

Overall, cross-sectionally, partial correlations observed between each of our three measures of cognition (ADAS-Cog-13, CDR-Sum Boxes, and AVLT-Total) and amyloid deposition, were significantly stronger when a cerebral white matter reference region was used after co-varying for age and education (Figure 1 and Table 2A: Overall Steiger’s Test p<1.39E-06). The strongest correlation was observed between ADAS-Cog-13 and FBP mcSUVR_wm_ (Figure 1 and Table 2A: Overall ADAS-Cog-13/FBP mcSUVR_cb_ r=0.43 (p=3.85E-51) and ADAS-Cog-13/FBP mcSUVR_wm_ r=0.54 (p=1.02E-148); Steiger’s Test p=7.63E-12).

**Figure 1:**
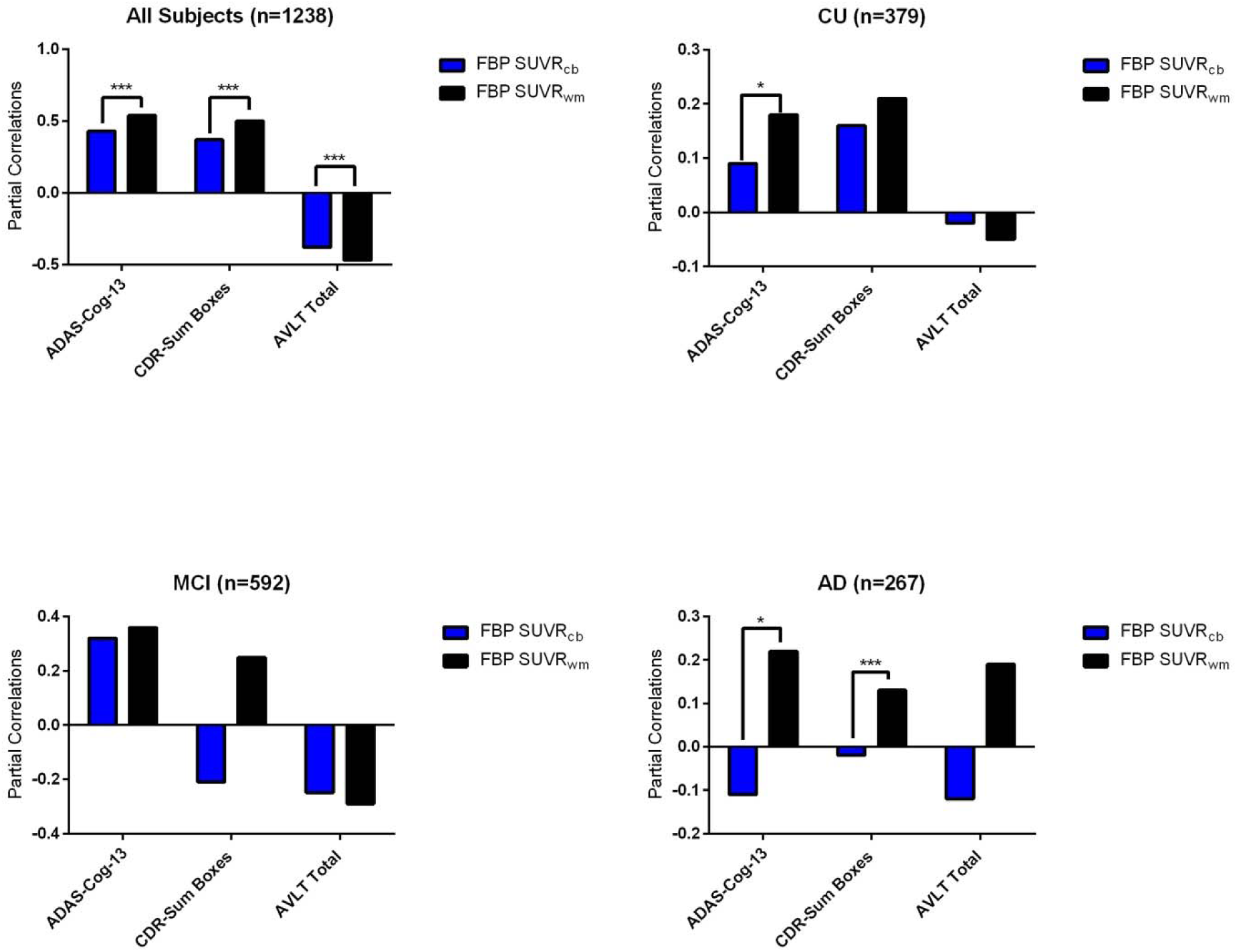
Partial Correlations between Cross Sectional Measurements of Memory Severity and Amyloid Deposition in the Overall, CU, MCI, and AD ADNI Groups: FBP mcSUVR_wm_ vs FBP mcSUVR_cb_

**Table 2:**
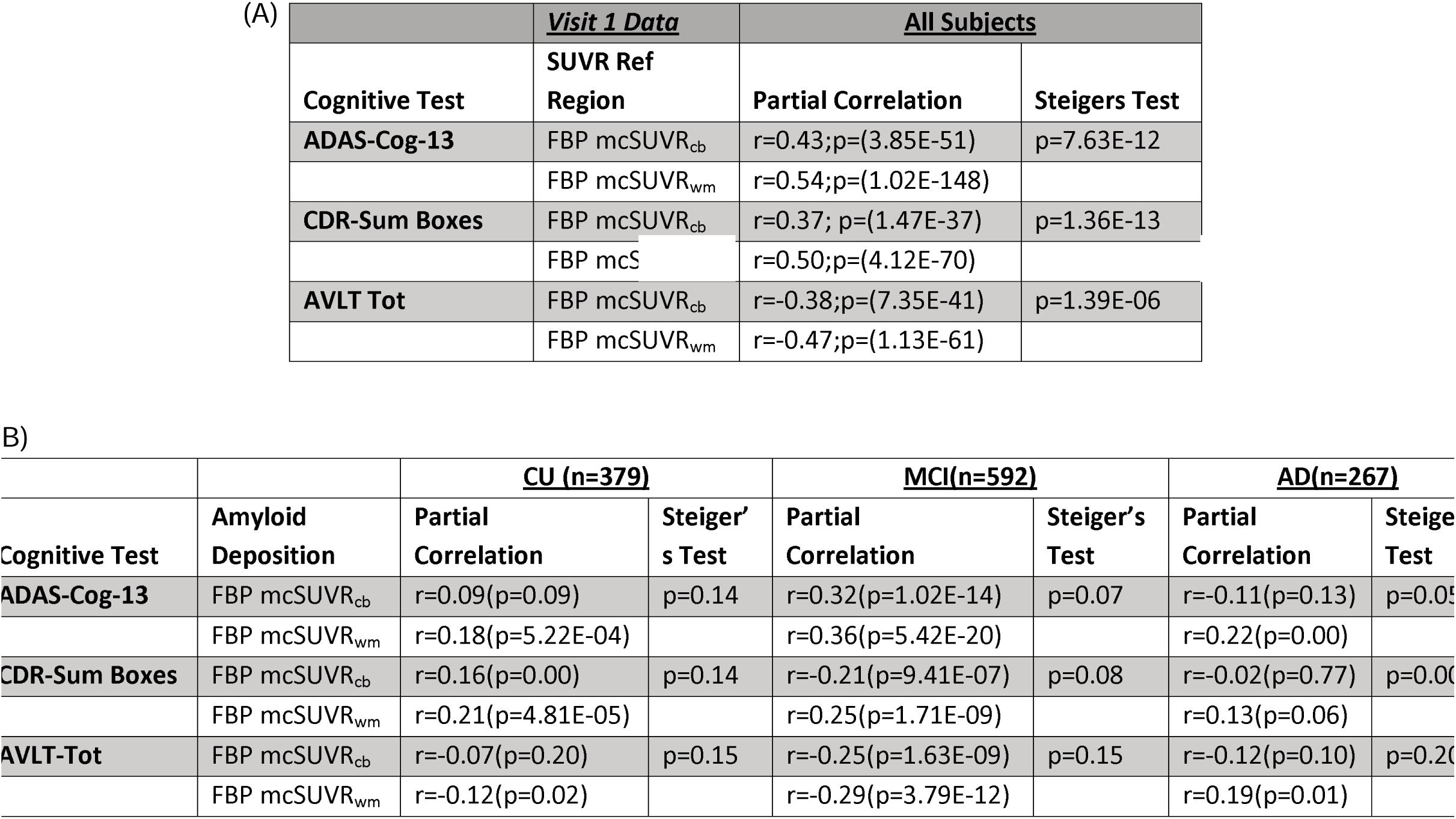
Partial Correlations between Cross Sectional Measurements of Clinical Seve Amyloid Deposition in the Overall Group (n=1238, **Table 2A) and in each CU, MCI MCI and AD subgroup (Table 2B): FBP mcSUVR_wm_ vs FBP mcSUVR_cb_. Data Associated with Figure 1**.

Longitudinally, greater partial correlations between cognition and FBP mcSUVR_wm_ were observed in the overall group (Figure 2 and Table 3A). Notably, the only significant partial correlations were observed when white matter was used as a reference region. No significant partial correlations were observed when a cerebellar reference region was used (Figure 2, Table 3A: Steiger’s Test p<0.00 all). The strongest longitudinal associations were observed between amyloid deposition using white matter as a reference region and CDR-Sum Boxes (Figure 2 and Table 3A: Overall CDR-Sum-Boxes/FBP mcSUVR_cb_ r=-0.10 (p=0.02) and CDR-Sum-Boxes/FBP mcSUVR_wm_ r=0.24 (p=7.85E-08), Steiger’s Test p=0.00).

**Figure 2:**
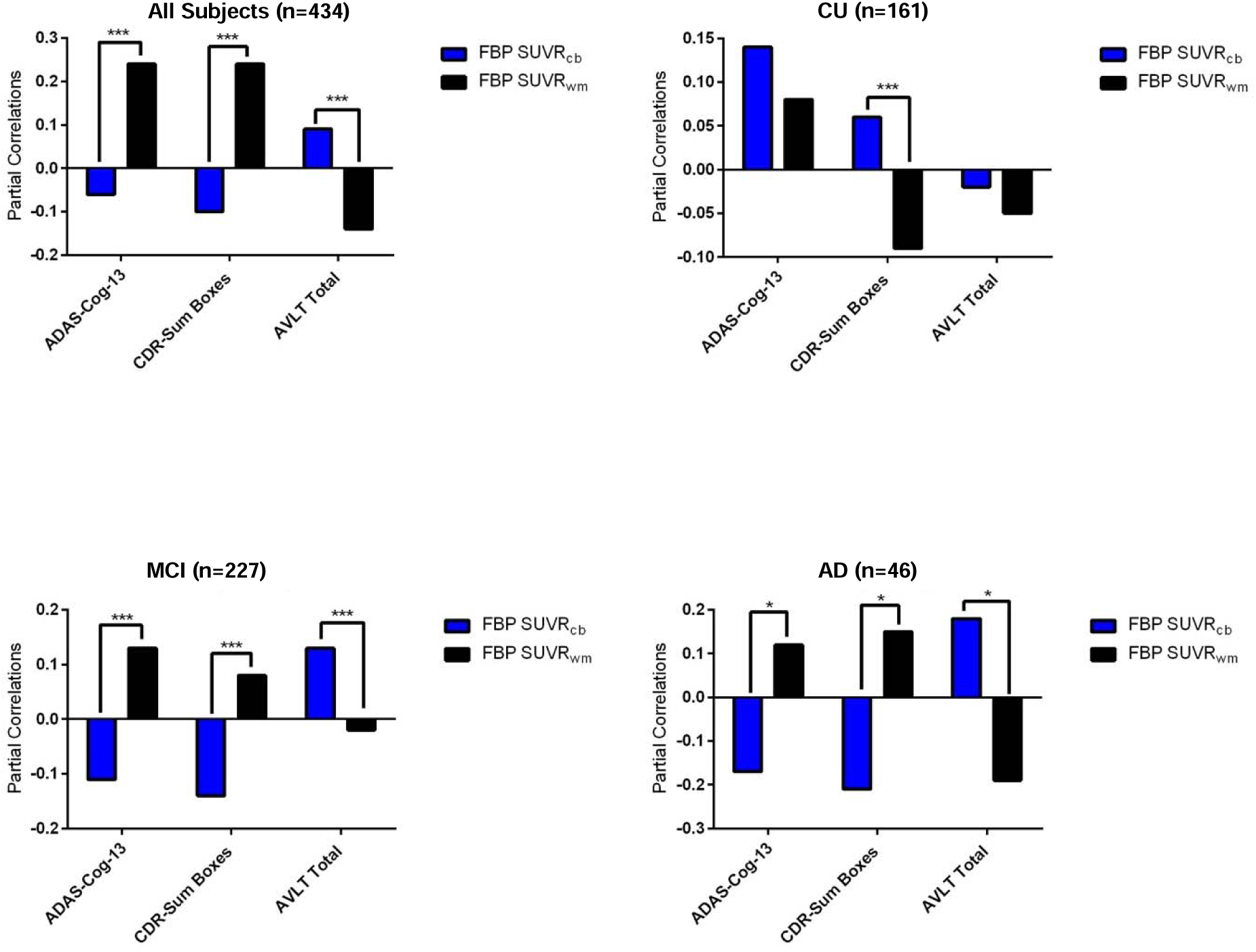
Partial Correlations between Longitudinal Measurements of Cognitive Decline and Amyloid Deposition in the Overall, CU, MCI, and AD ADNI Groups: FBP mcSUVR_wm_ vs FBP

**Table 3:** Partial Correlations between Longitudinal Measurements of Cognitive Decline and Amyloid Deposition in the Overall Groups (n=434, **Table 3A) and in each of the 3 sub-groups (Table 3B): FBP mcSUVR_wm_ vs FBP mcSUVR_cb_. Data Associated with Figure 2**.

|  |  | **All Subjects (n=434)** |  |
| --- | --- | --- | --- |
| **Cognitive Test** | **Amyloid Deposition** | **Partial Correlation** | **Steigers Test** |
| **ADAS-Cog-13** | FBP mcSUVR<sub>cb</sub> | r=-0.06(p=0.21) | p=0.00 |
|  | FBP mcSUVR<sub>wm</sub> | r=0.24(p=1.05E-07) |  |
| **CDR-Sum Boxes** | FBP mcSUVR<sub>cb</sub> | r=-0.10(p=0.02) | p=0.00 |
|  | FBP mcSUVR<sub>wm</sub> | r=0.24(p=7.85E-08) |  |
| **AVLT Tot** | FBP mcSUVR<sub>cb</sub> | r=0.09(p=0.05) | p=8.88e-16 |
|  | FBP mcSUVR<sub>wm</sub> | r=-0.14(p=0.00) |  |

(B)
|  |  | **CU (n=161)** |  | **MCI(n=227)** |  | **AD(n=46)** |  |
| --- | --- | --- | --- | --- | --- | --- | --- |
| **Cognitive Test** | **Amyloid Deposition** | **Partial Correlation** | **Steiger's Test** | **Partial Correlation** | **Steiger's Test** | **Partial Correlation** | **Steiger's Test** |
| **ADAS-Cog-13** | FBP mcSUVR<sub>cb</sub> | r=0.14(p=0.08) | p=0.33 | r=-0.11(p=0.10) | p=2.83E-08 | r=-0.17(p=0.28) | p=0.00 |
|  | FBP mcSUVR<sub>wm</sub> | r=0.08(p=0.30) |  | r=0.13(p=0.05) |  | r=0.12(p=0.43) |  |
| **CDR-SB** | FBP mcSUVR<sub>cb</sub> | r=0.06(p=0.43) | p=0.01 | r=-0.14(p=0.03) | p=4.80E-07 | r=-0.21(p=0.17) | p=0.00 |
|  | FBP mcSUVR<sub>wm</sub> | r=-0.09(p=0.26) |  | r=0.08(p=0.26) |  | r=0.15(p=0.32) |  |
| **AVLT-Tot** | FBP mcSUVR<sub>cb</sub> | r=-0.02(p=0.80) | p=0.64 | r=0.13(p=0.05) | p=3.98E-04 | r=0.18(p=0.25) | p=0.00 |
|  | FBP mcSUVR<sub>wm</sub> | r=-0.05(p=0.52) |  | r=-0.02(p=0.76) |  | r=-0.19(p=0.22) |  |

Post-hoc analysis was further conducted within AD continuum groups: CU, MCI, and AD. Cross-sectionally, the strongest and most significant correlations between amyloid deposition and memory/cognitive impairment tests were observed in the MCI group (Figure 1 and Table 2B: all r>0.21 (absolute value) (p<9.41E-07)). Within the CU and MCI group, significant associations between amyloid FBP mcSUVR and cognition were observed mostly when white matter was used as a reference region, although the difference between the partial correlations using white matter and cerebellum as a reference region were not significant (Example Table 2B: **CU:** ADAS-Cog-13/FBP mcSUVR_cb_ r=0.09(p-0.09), ADAS-Cog-13/FBP mcSUVR_wm_ r-0.18 (p=5.22E-04), Steiger’s Test (p=0.14); CDR-Sum Boxes/FBP mcSUVR_cb_ r=0.16(p-0.00), CDR-Sum Boxes/FBP mcSUVR_wm_ r=0.21 (p=4.81E-05)), Steiger’s Test (p=0.14); AVLT-Total/FBP mcSUVR_cb_ r=-0.07 (p=0.20) and AVLT-Total/FBP mcSUVR_wm_ r=-0.12 (p=0.02), Steiger’s Test p=0.15). Significant differences between FBP amyloid deposition values were only noted in the AD group (Table 2B: **AD:** Steiger’s Test: p=0.05 ADAS-Cog-13/FBP mcSUVR_cb_ r=-0.11(p=0.13) and ADAS-Cog-13/FBP mcSUVR_wm_ r=0.22(p=0.00) and Steiger’s Test: p=0.00 CDR-Sum Boxes/FBP mcSUVR_cb_ r=-0.02(p=0.77) and CDR-Sum Boxes/FBP mcSUVR_wm_ r=0.12(p=0.10) and Steiger’s Test: p= 0.20 AVLT-Tot/FBP mcSUVR_cb_ r=0.12 (p=0.10) and AVLT-Tot/FBP mcSUVR_wm_ r=0.19 (p=0.01)).

Longitudinally, over the whole group (Table 3A), it is interesting to note that we only observed significant correlations between ADAS-Cog-13 and amyloid accumulation when a cerebral white matter reference region was used (Table 3A: ADAS-Cog-13/FBP mcSUVR_cb_ r=-0.06(p=0.2) and ADAS-Cog-13/FBP mcSUVR_wm_ r=0.24(p=1.05E-07)). It is also interesting to note that the significant *negative* correlation of mcSUVR_cb_ with CDR-SB is paradoxical. For the sub-group post-hoc analysis (Table 3B), we noted non-significant correlation using either cerebral white matter or cerebellar reference region in each CU and in AD group separately used. For the AD group, such non-significance is partially due to small sample size and the ceiling effect on change in both amyloid accumulation and cognition. In the MCI group, we again observed paradoxical results such as the negative/positive correlation of CDR-SB/ALVT-Tol with amyloid when using cerebellar reference region. This is also true numerically for ADAS-Cog-13 (p=NS). For ADAS-Cog-13, the significant correlation with mcSUVR_wm_ is also significantly different from the non-significant correlation with mcSUVR_cb_ (Steiger’s Test: p=2.83E-08).

## Discussion

Here, we find that a white matter reference region is significantly better in characterizing the relationship between cross-sectional and longitudinal Florbetapir SUVR indicators of amyloid plaque burden and corresponding measurements of cognitive or clinical decline in an aggregate mild dementia, MCI and CU group. In a previous study, we found that a cerebral white matter reference region was significantly better than the traditional cerebellar reference region in detecting and tracking florbetapir SUVR increases in amyloid plaque deposition. Together these findings, support the potential use of a cerebral white matter reference region in the detection and tracking of amyloid plaque deposition using florbetapir PET, and suggest the need to clarify the generalizability of this conclusion to other amyloid PET ligands.

In our post-hoc within-group analysis, similar trends were observed cross-sectionally, particularly within the MCI group. Importantly, the overall group associations mentioned above could be driven by the associations observed within our MCI subgroup since most participants in this study were within this stage of AD. Longitudinally, few significant within-group associations were found between amyloid deposition and changes in clinical severity. However, out of the few significant associations noted, the only biologically sound associations (when amyloid deposition increases, clinical severity worsens) were demonstrated when a white matter reference ROI was used.

Amyloid beta accumulation is thought to be the initiating event in AD pathology triggering the formation of neurofibrillary tau tangles, neurodegeneration, and ultimately cognitive decline. The weak to moderate associations between amyloid deposition and cognition reported in literature is often attributed to the temporality of the amyloid cascade hypothesis. Since amyloid deposition occurs earlier in the disease process, amyloid is thought to be less significantly related to cognitive decline. Instead, the initial event of amyloid beta rise, subsequent tau accumulation, and the resulting sequence of amyloid and tau changes, particularly in areas such as the neocortex, is thought to mediate the association between initial amyloid deposition and cognitive decline – a finding supported by both post-mortem and in-vivo studies (Hanseeuw et al., 2019; Nelson et al., 2012).

However, our results suggest we may not be accurately capturing the strength of the relationship between amyloid accumulation and cognition at earlier stages of the AD continuum. In support of a stronger association, Jansen et al. showed a 10% higher prevalence of CSF-based amyloid abnormalities using CSF-based estimates compared to amyloid PET-based measurements in participants with normal cognition and mild cognitive impairment (MCI) (Jansen et al., 2022). Additionally, in a study using longitudinal data from amyloid negative individuals by Landau et al., the authors showed that baseline memory decline is associated with subthreshold amyloid accumulation (Landau et al., 2018). In both studies, the more significant associations observed in the **CU** (cognitively unimpaired) and **MCI** groups may reflect a stronger role of amyloid beta in these earlier stages of AD, where amyloid beta accumulation is more pronounced. In contrast, the weaker associations observed in **AD dementia** could be due to the introduction of other pathological players in AD such as tau deposition, and the plateauing of amyloid beta accumulation.

In comparison to the white matter reference region used in this study, a study in 2022 found that the whole cerebellum was an unstable reference region showing significant variation in FBP SUV values over time (Bourgeat et al., 2022). The same study further found that including white matter as a reference region could improve the harmonization between FBP and PiB, and the correlation between amyloid deposition and cognition, specifically MMSE (Bourgeat et al., 2022). In an abstract presented in 2016 to Human Amyloid Imaging Conference, Villemagne et al. additionally found that for FBP tracer, the white matter reference region demonstrated the least non-significant variation across time, diagnosis and amyloid status (Villemagne et al. 2016 abstract, HAI). Furthermore, in a study done by Lopez-Gonzales et al. in 2019, including white matter in the reference region improved semi-quantification of amyloid PET tracers by counteracting the effects of white matter signal spill into cortical target regions (López-González et al., 2019).

The current study has several limitations: First, this study has not yet explored the generalizability of findings regarding the value of using a white matter reference region to detect and track amyloid plaque deposition using other amyloid PET ligand-derived SUVRs. Additional studies are needed to address that issue and characterize and compare the potential use of centiloids in the detection and tracking of amyloid plaque deposition and the evaluation of amyloid plaque-modifying drugs in the treatment and prevention of AD. Second, our findings relied on the use of data from an aggregate group of dementia, MCI, and CU adults with and without amyloid plaque deposition. Cross-sectional and longitudinal studies involving many more research participants are needed to clarify the impact of using a cerebral white matter reference region on the association between amyloid plaque deposition on cognitive decline in dementia, MCI and CU sub-groups. Fourth, it remains possible that relationship between amyloid deposition and cognition could be influenced by other factors, such as tau deposition, vascular brain injury, neuroinflammation, etc. Finally, this study does not fully address the potentially confounding effects of age and AD-related-declines in white matter volumes, which could increase SUVRs using a white matter reference region due to the combined effects of atrophy and partial-volume averaging (Jagust et al 2023).

Overall, our findings support the further evaluation and potential of white matter reference region when SUVRs are used to detect and track amyloid plaque deposition and investigate effects of amyloid plaque-modifying treatments.

## Data Availability

All data used in this manuscript are available online at https://adni.loni.usc.edu/

## Disclosure Statement

No potential conflicts of interest relevant to this article exist.

## Acknowledgements

National Institute on Aging (NIA) grant P30AG072980, the Arizona Department of Health Services (ADHS) and the state of Arizona (ADHS Grant No. CTR057001). Research reported in this publication was also supported by the National Institute of Aging of the National Institutes of Health under award number T32AG082631.

## Notes

### Competing Interest Statement

The authors have declared no competing interest.

### Author Declarations

This study used data from the Alzheimer's Disease Neuroimaging Initiative which can be found at https://adni.loni.usc.edu/.

## Reference

Aizenstein, H. J., Nebes, R. D., Saxton, J. A., Price, J. C., Mathis, C. A., Tsopelas, N. D., Ziolko, S. K., James, J. A., Snitz, B. E., Houck, P. R., Bi, W., Cohen, A. D., Lopresti, B. J., DeKosky, S. T., Halligan, E. M., & Klunk, W. E. (2008). Frequent amyloid deposition without significant cognitive impairment among the elderly. Arch Neurol, 65(11), 1509–1517. 10.1001/archneur.65.11.1509

Berg, L. (1988). Clinical Dementia Rating (CDR). Psychopharmacol Bull, 24(4), 637–639.

Bourgeat, P., Doré, V., Burnham, S. C., Benzinger, T., Tosun, D., Li, S., Goyal, M., LaMontagne, P., Jin, L., Rowe, C. C., Weiner, M. W., Morris, J. C., Masters, C. L., Fripp, J., & Villemagne, V. L. (2022). β-amyloid PET harmonisation across longitudinal studies: Application to AIBL, ADNI and OASIS3. Neuroimage, 262, 119527. 10.1016/j.neuroimage.2022.119527

Chen, K., Roontiva, A., Thiyyagura, P., Lee, W., Liu, X., Ayutyanont, N., Protas, H., Luo, J. L., Bauer, R., Reschke, C., Bandy, D., Koeppe, R. A., Fleisher, A. S., Caselli, R. J., Landau, S., Jagust, W. J., Weiner, M. W., Reiman, E. M., & Alzheimer’s Disease Neuroimaging, I. (2015). Improved power for characterizing longitudinal amyloid-beta PET changes and evaluating amyloid-modifying treatments with a cerebral white matter reference region. J Nucl Med, 56(4), 560–566. 10.2967/jnumed.114.149732

Ciarmiello, A., Tartaglione, A., Giovannini, E., Riondato, M., Giovacchini, G., Ferrando, O., De Biasi, M., Passera, C., Carabelli, E., Mannironi, A., Foppiano, F., Alfano, B., & Mansi, L. (2019). Amyloid burden identifies neuropsychological phenotypes at increased risk of progression to Alzheimer’s disease in mild cognitive impairment patients. Eur J Nucl Med Mol Imaging, 46(2), 288–296. 10.1007/s00259-018-4149-2

Guillozet, A. L., Weintraub, S., Mash, D. C., & Mesulam, M. M. (2003). Neurofibrillary tangles, amyloid, and memory in aging and mild cognitive impairment. Arch Neurol, 60(5), 729–736. 10.1001/archneur.60.5.729

Hanseeuw, B. J., Betensky, R. A., Jacobs, H. I. L., Schultz, A. P., Sepulcre, J., Becker, J. A., Cosio, D. M. O., Farrell, M., Quiroz, Y. T., Mormino, E. C., Buckley, R. F., Papp, K. V., Amariglio, R. A., Dewachter, I., Ivanoiu, A., Huijbers, W., Hedden, T., Marshall, G. A., Chhatwal, J. P., … Johnson, K. (2019). Association of Amyloid and Tau With Cognition in Preclinical Alzheimer Disease: A Longitudinal Study. JAMA Neurol, 76(8), 915–924. 10.1001/jamaneurol.2019.1424

Iaccarino, L., Burnham, S. C., Tunali, I., Wang, J., Navitsky, M., Arora, A. K., & Pontecorvo, M. J. (2025). A practical overview of the use of amyloid-PET Centiloid values in clinical trials and research. Neuroimage Clin, 46, 103765. 10.1016/j.nicl.2025.103765

Jansen, W. J., Janssen, O., Tijms, B. M., Vos, S. J. B., Ossenkoppele, R., Visser, P. J., Aarsland, D., Alcolea, D., Altomare, D., von Arnim, C., Baiardi, S., Baldeiras, I., Barthel, H., Bateman, R. J., Van Berckel, B., Binette, A. P., Blennow, K., Boada, M., Boecker, H., … Zetterberg, H. (2022). Prevalence Estimates of Amyloid Abnormality Across the Alzheimer Disease Clinical Spectrum. JAMA Neurol, 79(3), 228–243. 10.1001/jamaneurol.2021.5216

Landau, S. M., Horng, A., & Jagust, W. J. (2018). Memory decline accompanies subthreshold amyloid accumulation. Neurology, 90(17), e1452–e1460. 10.1212/wnl.0000000000005354

López-González, F. J., Moscoso, A., Efthimiou, N., Fernández-Ferreiro, A., Piñeiro-Fiel, M., Archibald, S. J., Aguiar, P., & Silva-Rodríguez, J. (2019). Spill-in counts in the quantification of (18)F-florbetapir on Aβ-negative subjects: the effect of including white matter in the reference region. EJNMMI Phys, 6(1), 27. 10.1186/s40658-019-0258-7

Michalowska, M. M., Herholz, K., Hinz, R., Amadi, C., McInnes, L., Anton-Rodriguez, J. M., Karikari, T. K., Blennow, K., Zetterberg, H., Ashton, N. J., Pendleton, N., & Carter, S. F. (2022). Evaluation of in vivo staging of amyloid deposition in cognitively unimpaired elderly aged 78-94. Mol Psychiatry, 27(10), 4335–4342. 10.1038/s41380-022-01685-6

Nelson, P. T., Alafuzoff, I., Bigio, E. H., Bouras, C., Braak, H., Cairns, N. J., Castellani, R. J., Crain, B. J., Davies, P., Tredici, K. D., Duyckaerts, C., Frosch, M. P., Haroutunian, V., Hof, P. R., Hulette, C. M., Hyman, B. T., Iwatsubo, T., Jellinger, K. A., Jicha, G. A., … Beach, T. G. (2012). Correlation of Alzheimer Disease Neuropathologic Changes With Cognitive Status: A Review of the Literature. Journal of Neuropathology & Experimental Neurology, 71(5), 362–381. 10.1097/NEN.0b013e31825018f7

Pemberton, H. G., Collij, L. E., Heeman, F., Bollack, A., Shekari, M., Salvadó, G., Alves, I. L., Garcia, D. V., Battle, M., Buckley, C., Stephens, A. W., Bullich, S., Garibotto, V., Barkhof, F., Gispert, J. D., & Farrar, G. (2022). Quantification of amyloid PET for future clinical use: a state-of-the-art review. Eur J Nucl Med Mol Imaging, 49(10), 3508–3528. 10.1007/s00259-022-05784-y

Rosen, W. G., Mohs, R. C., & Davis, K. L. (1984). A new rating scale for Alzheimer’s disease. Am J Psychiatry, 141(11), 1356–1364. 10.1176/ajp.141.11.1356

Souto, J. J., Silva, G. M., Almeida, N. L., Shoshina, II, Santos, N. A., & Fernandes, T. P. (2021). Age-related episodic memory decline and the role of amyloid-β: a systematic review. Dement Neuropsychol, 15(3), 299–313. 10.1590/1980-57642021dn15-030002

Stevens, D. A., Workman, C. I., Kuwabara, H., Butters, M. A., Savonenko, A., Nassery, N., Gould, N., Kraut, M., Joo, J. H., Kilgore, J., Kamath, V., Holt, D. P., Dannals, R. F., Nandi, A., Onyike, C. U., & Smith, G. S. (2022). Regional amyloid correlates of cognitive performance in ageing and mild cognitive impairment. Brain Commun, 4(1), fcac016. 10.1093/braincomms/fcac016

Suppiah, S., Didier, M. A., & Vinjamuri, S. (2019). The Who, When, Why, and How of PET Amyloid Imaging in Management of Alzheimer’s Disease-Review of Literature and Interesting Images. Diagnostics (Basel), 9(2). 10.3390/diagnostics9020065

Villemagne, V. L., Burnham, S., Bourgeat, P., Brown, B., Ellis, K. A., Salvado, O., Szoeke, C., Macaulay, S. L., Martins, R., Maruff, P., Ames, D., Rowe, C. C., & Masters, C. L. (2013). Amyloid β deposition, neurodegeneration, and cognitive decline in sporadic Alzheimer’s disease: a prospective cohort study. Lancet Neurol, 12(4), 357–367. 10.1016/s1474-4422(13)70044-9

Villemagne, V. L., Pike, K. E., Chételat, G., Ellis, K. A., Mulligan, R. S., Bourgeat, P., Ackermann, U., Jones, G., Szoeke, C., Salvado, O., Martins, R., O’Keefe, G., Mathis, C. A., Klunk, W. E., Ames, D., Masters, C. L., & Rowe, C. C. (2011). Longitudinal assessment of Aβ and cognition in aging and Alzheimer disease. Ann Neurol, 69(1), 181–192. 10.1002/ana.22248

